# Coding of No Drug Use at Discharge in Public-Use Treatment Data and Federal Performance Tables

**DOI:** 10.64898/2026.11.25.26368966

**Authors:** Lin Wang, Jennifer L. Brown, Dayu Sun, Nan Miles Xi

## Abstract

Drug use at discharge from substance use treatment is a national outcome measure that the Substance Abuse and Mental Health Services Administration computes for state block grant performance tables. Researchers analyze it in the public-use files of the Treatment Episode Data Set Discharges (TEDS-D). The federal specification codes the frequency of use for a “None” substance as not applicable. We examined how the TEDS-D public-use files for 2015 to 2024 code these records. From 2018 to 2024, every public-use record with “None” as the primary discharge substance had its frequency coded as missing or unknown. We call such records with no other discharge substance all-None records. All-None records were 12.5% of discharges in 2024. Among 729,346 discharges from 2018 to 2024 of adults admitted with daily use of heroin, other opioids, methamphetamine, or cocaine who stayed at least 31 days and did not die in treatment, 64,821 (8.9%) were all-None records. Massachusetts and New Jersey, whose discharge forms document “None” as reported no use, contributed 81.7% of them. With all-None records dropped, New Jersey’s rate of no use at discharge was 6.8% (49th of 51 jurisdictions). Counting them as no use raised it to 52% (18th). A 2021 reporting change in Massachusetts implied that 98% (95% CI, 97% to 99%) of its all-None records reported no use. In 2024, New Jersey’s block grant table reported 98.9% abstinence at discharge in short-term residential treatment among clients who used drugs at admission. The public-use frequency field gave 14.7%. Counting “None” as no use reproduced the federal values within 4.1 percentage points in every setting and year. A published national analysis excluded 93.9% of completed episodes in New Jersey. Of these, 95.2% were all-None records. Analyses that define use from the frequency field drop documented reports of no use in New Jersey and Massachusetts.

## Introduction

The Treatment Episode Data Set Discharges (TEDS-D) compiles state reports of discharges from substance use treatment. Drug use at discharge is a national outcome measure of the Substance Abuse and Mental Health Services Administration (SAMHSA).^1^ SAMHSA computes it for the performance tables of state block grant reports.^2^ Researchers analyze it in the TEDS-D public-use files.^3^ The federal specification codes the frequency of use for a “None” substance as not applicable, separately from unknown.^1,4^ We examined how the public-use files code these records and how this affects state measures and research.

## Methods

We used the TEDS-D public-use files for 2015 to 2024.^3^ The main population comprised discharges from 2018 to 2024 of adults admitted with daily use of heroin, other opioids, methamphetamine, or cocaine as the primary substance. Stays lasted at least 31 days and did not end in death. Use at discharge was the reported frequency for the admission primary substance. We call discharge records with “None” as the primary discharge substance and no substance in the other fields all-None records.

We reviewed the discharge forms of Massachusetts and New Jersey^5,6^ and the 2020 and 2022 crosswalks of the 8 jurisdictions with the most all-None records.^7^ From the Massachusetts reporting change in 2021, we estimated the share of no use among all-None records (eMethods in Supplement 1).

For jurisdictions with at least 100 discharges of known use, we computed rates of no use at discharge with Wilson 95% CIs and ranks. All-None records were either dropped, as the public-use frequency field implies, or counted as no use in Massachusetts and New Jersey, whose forms document “None” as reported no use.

We reproduced New Jersey’s block grant Table 18, drug abstinence at discharge among clients using drugs at admission, for 2022 to 2024.^2^ We counted “None” as no use or used frequency fields only (eMethods in Supplement 1). We also reconstructed the sample of a national analysis of completed episodes from 2015 to 2019.^8^

Analyses used Python, version 3.12, in November 2026. This cross-sectional study of deidentified public data was not human participants research under the Common Rule (45 CFR §46.102) and needed no institutional review board review or informed consent. We followed the Strengthening the Reporting of Observational Studies in Epidemiology (STROBE) reporting guideline.^9^

## Results

From 2018 to 2024, every public-use discharge record with “None” as the primary substance had its frequency coded as missing, unknown, not collected, or invalid (160,431 of 160,431 in 2024). The codebook lists no not-applicable category. All-None records were 12.5% of discharges in 2024.

The main population included 729,346 discharges, of which 64,821 (8.9%) were all-None records. Massachusetts and New Jersey contributed 81.7% of them (Table). With all-None records dropped, the Massachusetts rate of no use fell from 88% in 2020 to 10% in 2021. Counting them as no use kept it between 86% and 88% in every year. The implied share of no use among all-None records was 98% (95% CI, 97%-99%).

**Table.** Documentation and Consequences of All-None Discharge Records in Massachusetts and New Jersey.

| **Characteristic** | **Massachusetts** | **New Jersey** |
|---|---|---|
| Discharge question in state forms | “Has there been any drug or alcohol use in the last 30 days or since Enrollment if in treatment less than 30 days?” Substance items are skipped after “No.”<sup>a</sup> | “Is client using drugs or alcohol at discharge?” Answers include “No Alcohol & No Drugs” and, separately, “Unknown.”<sup>b</sup> |
| State crosswalk, 2020 and 2022<sup>c</sup> | State code 0 “None” maps to “None.” Unknown, missing, invalid, and refused codes map to unknown. | “No Alcohol & No Drugs” maps to “None” and to no use in the past month. “Unknown” maps to not collected (substance) and unknown (frequency). |
| All-None records, No. (%)<sup>d</sup> | 26,609 (42.1) | 26,352 (34.4) |
| All-None records, % of discharges by year<sup>e</sup> | <1 in 2015-2020. 65-69 in 2021-2024. | 32-38 in every year, 2015-2024. |
| No use at discharge, all-None records dropped, % (95% CI) [rank]<sup>d</sup> | 73.2 (72.6-73.7) [6] | 6.8 (6.6-7.1) [49] |
| No use at discharge, all-None records counted as no use, % (95% CI) [rank]<sup>d</sup> | 87.4 (87.1-87.7) [2] | 52 (51.6-52.4) [18] |
| Evidence from a reporting change | 2021 change: implied share of no use among all-None records, 98% (95% CI, 97%-99%) | No reporting change in 2015-2024 |
| Federal block grant table, short-term residential treatment, 2024<sup>f</sup> | Not available<sup>g</sup> | 98.9% abstinent at discharge. Public-use frequency field: 14.7% (34 of 232). Only 376 of 8,527 discharges had a known frequency. |

<sup>a</sup> Residential Treatment Disenrollment Assessment Manual, version 14 (2017).<sup>5</sup>
<sup>b</sup> New Jersey Substance Abuse Monitoring System discharge questionnaire.<sup>6</sup>
<sup>c</sup> State data crosswalks filed with SAMHSA.<sup>7</sup>
<sup>d</sup> Main population: adults admitted with daily use of heroin, other opioids, methamphetamine, or cocaine who stayed at least 31 days and did not die in treatment, 2018 to 2024. Ranks are among 51 jurisdictions with at least 100 discharges of known use.
<sup>e</sup> Main population criteria applied to discharges from 2015 to 2024.
<sup>f</sup> Clients who used drugs at admission, excluding clients with planned medication for opioid use disorder, deaths, and incarcerations, as defined in the table notes.<sup>2</sup>
<sup>g</sup> We did not find a public Massachusetts block grant report.

With all-None records dropped, New Jersey’s rate of no use at discharge was 6.8% (95% CI, 6.6%-7.1%), ranking 49th of 51 jurisdictions. Counting them as no use raised it to 52% (95% CI, 51.6%-52.4%), ranking 18th (Figure, A). Massachusetts moved from 73.2% (6th) to 87.4% (2nd). No other jurisdiction moved by more than 1 rank. With any primary substance and any length of stay, New Jersey moved from 39th to 2nd of 52. In New Jersey, 71.6% of all-None records ended with treatment completion, compared with 13.8% of records with known use.

**Figure.**
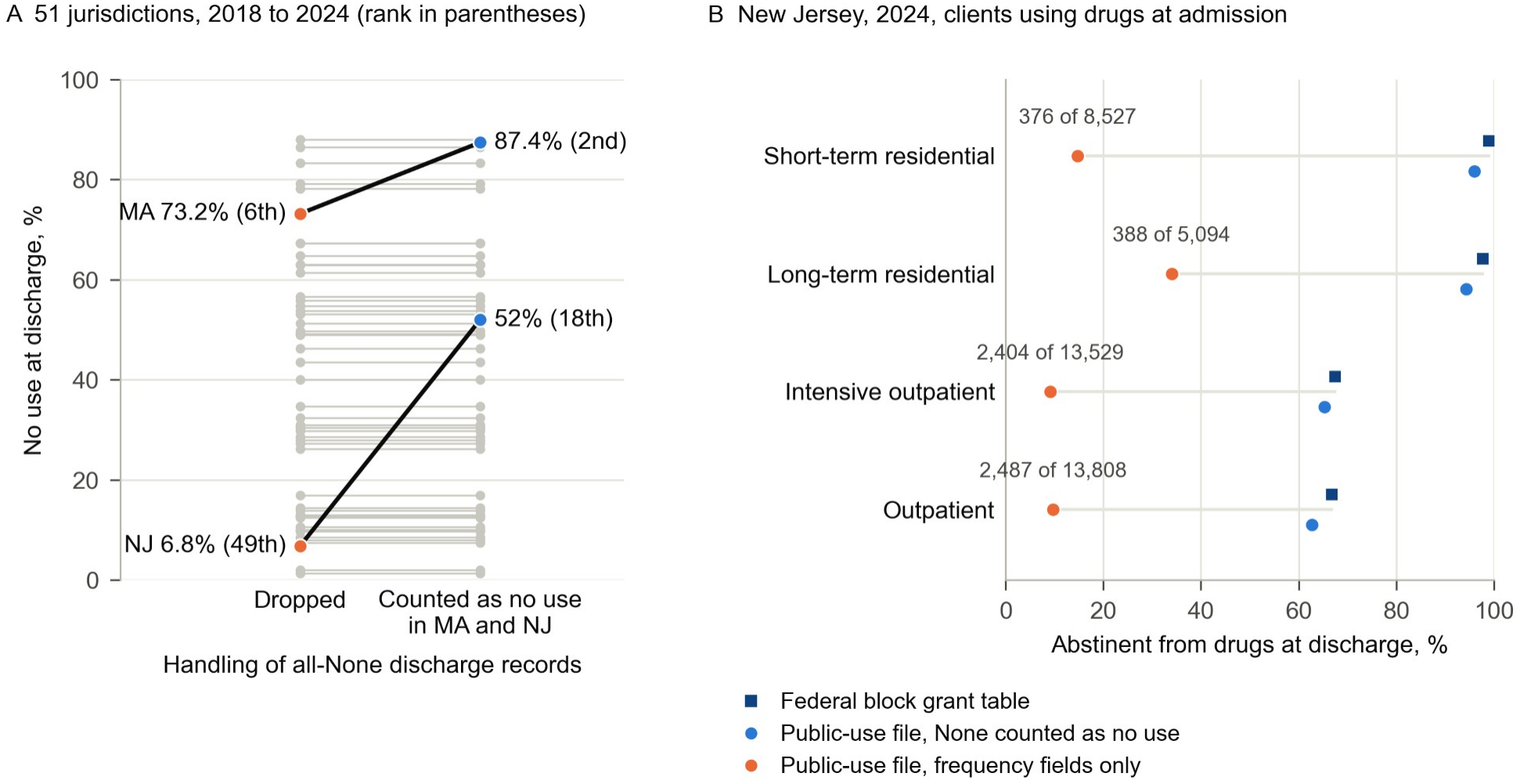
Rates of No Use at Discharge Under 2 Conventions for All-None Records and in New Jersey’s Federal Block Grant Table. A, Rate of no use at discharge in 51 jurisdictions, 2018 to 2024. The population comprised adults admitted with daily use of heroin, other opioids, methamphetamine, or cocaine who stayed at least 31 days and did not die in treatment. All-None records were dropped or counted as no use in Massachusetts (MA) and New Jersey (NJ). Gray lines show the other jurisdictions. Ranks among the 51 jurisdictions are in parentheses. B, Percentage of New Jersey clients in 2024 who used drugs at admission and were abstinent from drugs at discharge, by treatment setting. Values come from the federal block grant table and from the public-use files, with “None” counted as no use or with frequency fields only. Numbers above the frequency-only points give the discharges with a known frequency and all discharges in the table population.

The federal specification for this table counts a “None” substance with a not-applicable frequency as abstinence.^1^ In 2024, New Jersey’s table reported 98.9% abstinence at discharge among clients in short-term residential treatment who used drugs at admission (Figure, B). The public-use frequency field gave 14.7%. Counting “None” as no use reproduced the federal values within 4.1 percentage points in every setting and year.

Our reconstruction of the national analysis^8^ reproduced its 943,507 exclusions, which included 93.9% of New Jersey’s completed episodes. Of these, 95.2% were all-None records.

## Discussion

Two federal data products treat the same discharge records differently. The block grant tables count them as abstinent.^1^ The public-use files code their frequency as missing. Analyses that define use from the frequency field therefore drop documented reports of no use in New Jersey and Massachusetts.

Published studies have dropped these records^8^ or counted “None” as abstinence.^10^ Neither convention fits every state. In the 2022 Kansas crosswalk, for example, “None” also covers missing values.^7^ SAMHSA could keep the not-applicable code in the public-use files. Analysts could then follow each state’s crosswalk and report their convention.

Limitations include discharge forms for only 2 states, a shorter recall window in New Jersey, an untestable assumption behind the Massachusetts estimate, and approximate reproduction of the federal table.

## Supporting information

Supplement 1 (eMethods)

Supplement 2 (Data Sharing Statement)

## Data Availability

The public-use files of the Treatment Episode Data Set Discharges are available from the Substance Abuse and Mental Health Services Administration at https://www.samhsa.gov/data/data-we-collect/teds-treatment-episode-data-set/datafiles. The New Jersey block grant reports and the 2020 and 2022 state data crosswalks are public, with URLs in the references. The analysis code and aggregate results will be deposited in Zenodo at publication. Until then, the corresponding author will provide them on request.

## Article Information

### Author Contributions

Dr Wang had full access to all of the data in the study and takes responsibility for the integrity of the data and the accuracy of the data analysis.

### Concept and design

Wang, Brown.

### Acquisition, analysis, or interpretation of data

Wang, Brown, Sun, Xi.

### Drafting of the manuscript

Wang.

### Critical review of the manuscript for important intellectual content

Wang, Brown, Sun, Xi.

### Statistical analysis

Wang, Sun, Xi.

## Data Sharing Statement

See Supplement 2.

