## Supplement 1 (eMethods) for "Coding of No Drug Use at Discharge in Public-Use Treatment Data and Federal Performance Tables"

**eMethods.** Data, Definitions, and Reproduction of the Federal Block Grant Table

**eReferences**

This supplementary material has been provided by the authors to give readers additional information about their work.

### eMethods. Data, Definitions, and Reproduction of the Federal Block Grant Table

#### Data and Main Population

We used the Treatment Episode Data Set Discharges (TEDS-D) public-use files for discharge years 2015 to 2024.[1] The main population covered discharges from 2018 to 2024. It comprised adults (age 18 years or older) whose primary substance at admission was heroin, other opioids (TEDS category: other opiates and synthetics), methamphetamine, or cocaine (including crack). They used this substance daily in the month before admission. Stays lasted at least 31 days and did not end in death. Of 10,285,040 discharges from 2018 to 2024, 10,050,200 were of adults. Of these, 4,693,542 had 1 of the 4 primary substances. Daily use at admission was reported for 2,225,184 of them. Excluding deaths left 2,216,908 discharges, of which 729,346 lasted at least 31 days.

#### Use at Discharge and All-None Records

We searched the 3 discharge substance fields for the admission primary substance. A record had known use when that substance appeared with a frequency of no use in the past month, some use, or daily use, with the same value wherever it appeared. A record was an all-None record when "None" was the primary discharge substance and no other discharge field held a substance. In every public-use file from 2018 to 2024, the discharge frequency was coded −9 (missing, unknown, not collected, or invalid) whenever the primary discharge substance was "None." The federal specifications code this frequency as not applicable, separately from unknown and not collected.[2,3] The codebook lists no not-applicable category.[1]

#### State Rates and Ranks

For each jurisdiction with at least 100 discharges of known use, the rate of no use at discharge was the share with no use among discharges of known use. We call this dropping all-None records. Counting added the all-None records of Massachusetts and New Jersey to the numerator and the denominator because the discharge forms of both states document "None" as reported no use. We computed Wilson 95% CIs and ranked jurisdictions from the highest rate. No 2 jurisdictions had the same rate. Fifty-one jurisdictions met the threshold: 49 states, the District of Columbia, and Puerto Rico. New Mexico had fewer than 100 discharges of known use.

If all-None records were counted as no use in every jurisdiction, New Jersey would rank 20th and Massachusetts 2nd. Kansas would move from 9th to 3rd, Puerto Rico from 46th to 37th, and Arizona from 39th to 36th. In a broader population of adults with any primary substance used in the month before admission and any length of stay, New Jersey moved from 10.7% (39th of 52 jurisdictions) to 80.8% (2nd).

#### Massachusetts Reporting Change

Among discharges meeting the main population criteria, all-None records made up less than 1% of Massachusetts discharges from 2015 to 2020 and 65% to 69% from 2021 to 2024. The Massachusetts crosswalks for 2020 and 2022 map "None" in the same way and do not explain the change. We compared a low window (2018 to 2020) with a high window (2021 to 2023, the first 3 years after the change). Within each drug, the implied composition of all-None records in the high window was

$$\varepsilon_g = [f_{g,L} - f_{g,H} + (o_L - o_H)\ \pi_{g,L}] \ / \ e_H,$$

where $f_g$ is the share of all records in use group $g$ (no use, some use, or daily use) and $o$ is the share of other records with unknown use. The subscripts L and H mark the low and high windows. The composition of records with known use in the low window is $\pi_{g,L}$. The share of all-None records in the high window is $e_H$. The formula assumes that the distribution of use within each drug did not change between the windows. It also assumes that other records with unknown use had the known composition of the low window.

We reweighted the low window to the covariate distribution of the high window with a logistic model for window membership. The covariates were age group, sex, race and ethnicity, service setting, referral source, prior treatment, employment, living arrangement, arrests, psychiatric problem, and drug. Race and ethnicity were reported by treatment facilities in fixed TEDS-D categories. We included them because the client mix differed between the windows. Drug-specific estimates were combined in proportion to their numbers of all-None records. Percentile 95% CIs came from 200 bootstrap samples drawn within windows. The reweighted share of no use among all-None records was 98% (95% CI, 97%-99%). The unweighted estimate was 101% (95% CI, 100%-101%). The estimator is not bounded by 100%. The unweighted version also absorbs the change in client mix between the windows.

**Reproduction of the Federal Block Grant Table**

Table 18 of New Jersey's block grant reports for fiscal years 2024 to 2026 gives drug abstinence at discharge for calendar years 2022 to 2024, by treatment setting (level of care).[4-6] It reports all clients and clients using drugs at admission. Following the federal specification for this table[3] and the table notes, we kept New Jersey discharges from short-term residential, long-term residential, intensive outpatient, and outpatient care. We excluded clients with planned medication for opioid use disorder, deaths, and incarcerations. Drugs were all substances other than alcohol. Use meant some or daily use.

At each time point, a record was known when it reported drug use. A record without reported drug use was known under the counting rule when 3 conditions held. Its primary substance was reported. At least 1 substance field was informative (a drug or alcohol with a known frequency, or "None" in the primary field). No reported drug had a missing frequency. This rule follows the federal specification, which counts a "None" substance with a not-applicable frequency as abstinence.[3] Counting "None" in any substance field gave values up to 17 percentage points above the federal values in outpatient settings. Under the frequency-only rule, a record without reported drug use was known when at least 1 frequency field was reported and no reported drug had a missing frequency. Records entered the comparison when they were known at both admission and discharge.

Across the 4 settings and 3 years, the counting rule differed from the federal percentages by −4 to 3.9 percentage points. The frequency-only rule gave 8% to 34%, compared with 56.9% to 99.5% in the federal table.

For each setting and year, let $E$ be the federal denominator for all clients and $A$ the federal number of clients abstinent at discharge. Let $M$ be the number of public-use records after exclusions that were not all-None records. Let $X$ be the number by which the federal count after exclusions exceeded the public-use count, or 0 if it did not. At least $E - (M + X)$ all-None records then entered the federal denominator. At least $A - (M + X)$ of them were counted as abstinent. In short-term residential care in 2024, at least 7,646 of 7,914 all-None records (96.6%) were counted as abstinent. Because the public-use files swap some records between states, this bound is approximate.

**Reconstructed National Sample**

We kept completed treatment episodes from the 2015 to 2019 files with no age restriction, as in the national analysis.[7] Of 3,354,858 completed episodes, 943,507 (28.1%) lacked a discharge frequency, the number of exclusions the analysis reported. In New Jersey, 188,937 of 201,164 completed episodes (93.9%) lacked a discharge frequency. Of these, 179,888 (95.2%) were all-None records.

**State Crosswalks**

We read the 2020 and 2022 crosswalks of the 8 jurisdictions with the most all-None records in the main population: Massachusetts, New Jersey, California, Arizona, Ohio, Kansas, Florida, and Puerto Rico.[8] The Kansas substance use crosswalk was available for 2022 only. The California file in the 2020 package is dated 2019. New Jersey maps the answer "No Alcohol & No Drugs" to "None" and to no use in the past month. Massachusetts maps its own "None" code to "None" and codes unknown values separately. The Massachusetts discharge forms we reviewed cover residential and acute care. Outpatient care accounted for 58% of the state's all-None records. California and Ohio map their own "None" codes to "None." Arizona defines "None" as no use in the past 30 days. Kansas maps missing substance values to "None." Florida maps "Presenting At-Risk" to "None." Puerto Rico's 2020 crosswalk mapped tobacco use to "None."

**Software and Confidentiality**

Analyses used Python, version 3.12.7 (Anaconda distribution), with pandas 2.2.2, NumPy 1.26.4, scikit-learn 1.5.1 for the reweighting model, and pyarrow 16.1.0. Figures were drawn with matplotlib, version 3.10.8. To protect confidentiality, the public-use files swap some records with records in another state.[1] The swap partner is sought first within the same census division, then outside the division, and then outside the region. State counts in the public-use files can therefore differ slightly from tables that SAMHSA computes from the full data.
