## Supplement 2 (Data Sharing Statement) for "Coding of No Drug Use at Discharge in Public-Use Treatment Data and Federal Performance Tables"

**Data available:** Yes.

**Data types:** Publicly available deidentified data and aggregate results.

**How to access data:** The Treatment Episode Data Set Discharges public-use files are available from the Substance Abuse and Mental Health Services Administration (SAMHSA) at https://www.samhsa.gov/data/data-we-collect/teds-treatment-episode-data-set/datafiles. The New Jersey block grant reports are listed as eReferences 4 to 6 in Supplement 1. The 2020 and 2022 state data crosswalks are listed as reference 7 of the article. The aggregate results will be deposited in Zenodo with the analysis code.

**When available:** The source data are available now. The aggregate results will be available with publication.

**Supporting Documents**

**Document types:** Statistical and analytic code.

**How to access documents:** The analysis code will be deposited in Zenodo, with a DOI provided at publication. Until then, the corresponding author will provide it on request.

**When available:** With publication.

**Additional Information**

**Who is providing access to the data:** SAMHSA provides the source data. The authors provide the code and aggregate results.

**Who can access the data:** Anyone. Use of the public-use files is subject to the data use terms of SAMHSA.

**Types of analyses:** Any.

**Mechanisms of data availability:** Public download.
